# Regional anaesthesia for early rib fracture associated pain management: Systematic review and network meta-analysis protocol

**DOI:** 10.1101/2025.03.17.25324085

**Authors:** Christopher Partyka, Scott Farenden, David Tian, Anthony Delaney, Kate Curtis

## Abstract

**Introduction:** Rib fractures result in significant pain that can impair ventilatory function and result in increased rates of respiratory complication. Early effective analgesia is a crucial treatment goal for these patients however, this is often difficult to achieve. Regional anaesthesia techniques have been used as analgesic adjuncts following thoracic trauma to reduce both pain scores and opioid consumption. More recently, the serratus anterior plane block and erector spinae block have been used in the field of Emergency Medicine as an early analgesic adjunct through either a single depot infiltration of local anaesthetic (‘single-shot’) or continuous infusion catheter-based blocks in patients with rib fractures. These single-shot blocks are relatively quick and logistically easy to perform. Within the field of Emergency Medicine, the limitations to performing invasive procedures (like catheter-insertion) under strict sterile conditions within an appropriate clinical space makes these single-shot techniques a much more viable option. We will conduct a systematic review of randomised clinical trials (RCTs) with network meta-analysis to test the effectiveness of these ‘single-shot’ regional anaesthesia techniques on early pain reduction in adult patients with rib fractures.

**Methods and analysis:** We will include RCTs that compare a single-shot regional anaesthesia technique for the purpose of rib fracture management to either placebo, a passive comparison (e.g. standard care management with analgesia), or an active comparison (e.g. one block against another – head-to-head trials). We will perform a search that includes the electronic databases PubMed, MEDLINE, EMBASE, CinAHL and CENTRAL plus clinical trial registries. Two reviewers will independently screen titles and abstracts, perform full article reviews and extract study data, with discrepancies resolved by a third reviewer. We will report study characteristics and quantify risk of bias. We will perform a traditional random-effects meta-analysis for each pairwise comparison and if there is sufficient data, network meta-analysis will be conducted using a Bayesian approach to simultaneously compare multiple treatments via a common comparator (standard care). We will evaluate overall certainty of evidence using the Grading of Recommendations Assessment, Development, and Evaluation framework.

**Conclusion:** This systematic review and network meta-analysis will provide clinicians with an overview of the effectiveness of the ‘single-shot’ regional anaesthesia techniques used for early pain reduction in adult patients with rib fractures.

## INTRODUCTION

Rib fractures resulting from blunt thoracic trauma are common injuries requiring admission to hospital which are associated with in-hospital mortality and a variety of complications^1^. The significant pain resulting from these fractures can impair ventilatory function and result in increased rates of nosocomial pneumonia, atelectasis, respiratory failure, and mortality^2^. Early effective analgesia is therefore a crucial treatment goal for these patients however, this is often difficult to achieve^2^.

Protocolized rib-fracture care bundles include the early implementation of multimodal analgesia combined with ventilatory support, physiotherapy, early mobilisation, and pain team consultation^3^. They have been demonstrated to reduce pain scores and the incidence of pneumonia in patients with rib fractures^4-6^ and form the current standard of care for these patients being admitted to hospital. Opiate-based patient-controlled opiate analgesia (PCA) is frequently used in these protocols. In the elderly, these drugs are associated with delirium, constipation, and falls^7,8^.

In addition to pain scores, respiratory function has become a physiological marker of interest in patients with rib fractures. Respiratory function can be measured by a variety of methods including traditional peak flow and spirometry. Given the inverse relationship between vital capacity and the development of respiratory complications^9^, bedside incentive spirometry has featured more regularly as a recorded outcome measure in rib fracture management research^10,11^. In the geriatric population, spirometry appears to predict outcomes and suitability for discharge with greater accuracy than pain scores^12^. More recently, diaphragmatic excursion measured by bedside ultrasound, has become a parameter of interest in the Intensive Care clinical setting^13,14^ and is also featuring in rib fracture management research^15^.

Regional anaesthesia techniques provide analgesia to most of the hemithorax by way of ultrasound-guided infiltration of local anaesthetic solution to specific nerves or tissue planes. They have been used as analgesic adjuncts following cardiothoracic and chest wall surgery to reduce both pain scores and opioid consumption^16,17^. Whilst thoracic regional anaesthesia techniques have been studied in detail for their role in perioperative analgesia, it is highly likely that perioperative studies for these techniques do not extrapolate well to the field of thoracic trauma. It is well described that tissue plane disruption resulting from rib fractures facilitate deeper and more posterior spread of local anaesthetic injections compared to a hemithorax with intact ribs^18^. More recently, the serratus anterior plane block (SAPB) and erector spinae block (ESB) have been used in the field of Emergency Medicine as an analgesic adjunct in treating rib fracture associated pain^19,20^. These blocks lead to a reduction in pain scores and opiate consumption following either a single depot infiltration of local anaesthetic (hereafter referred to as a ‘single-shot’) or continuous infusion catheter-based blocks in patients with rib fractures^15,21-24^. The single-shot fascial plane blocks are relatively quick and logistically easy to perform^25^. They provide clinicians who are frequently time-poor with multiple competing priorities, with a pragmatically efficient and safe procedure to offer patients. Within the field of Emergency Medicine, the limitations to performing invasive procedures (like catheter-insertion) under strict sterile conditions within an appropriate clinical space makes these single-shot techniques a much more viable option.

There have been several clinical trials investigating these blocks either in isolation (against a control) or in a head-to-head manner. The completed reviews (both systematic and scoping) have evaluated these techniques on an individual basis^1,16,26-28^. Network meta-analyses provide a technique for comparing multiple treatments simultaneously in a single analysis by combining direct and indirect evidence within a network of randomized controlled trials^29^. A recent comprehensive network meta-analysis and systematic review of regional anaesthesia modalities for blunt thoracic trauma demonstrated that thoracic epidurals showed efficacy in reducing pain at 24 hours, mechanical ventilation duration, and ICU and hospital stays^30^. The thoracic epidural technique, and many catheter-based blocks are not regularly available or feasible to perform in the Emergency Department during the hyperacute phases of care. This study will complete a systematic review and network meta-analysis of the ‘single-shot’ regional anaesthesia techniques used in the early management of rib fractures and report their measured treatment effects on pain relief and other outcomes.

We plan to conduct a systematic review of randomised clinical trials with network meta-analysis to test the effectiveness of ‘single-shot’ regional anaesthesia techniques on early pain reduction in adult patients with rib fractures.

## METHODS

We will conduct a systematic review and network meta-analysis in accordance with the PRISMA-P guidelines^31^. It has been registered at the international prospective register of systematic reviews (PROSPERO) (CRD420251003934). The review will be reported in accordance with the PRISMA (Preferred Reporting Items for Systematic Reviews and Meta-Analyses) 2020 and PRISMA Extension Statement for Reporting of Systematic Reviews Incorporating Network Meta-analyses of Health Care Interventions: Checklist and Explanations (PRISMA-NMA) checklists^32,33^.

### Eligibility criteria

#### Population

Studies will be included in this systematic review if they include patients undergoing treatment for rib fractures following thoracic trauma.

#### Intervention

The intervention is the administration of a single-shot regional anaesthesia technique for the purpose of rib fracture management.

Techniques will include serratus anterior plane block (SAPB), erector spinae block (ESB), paravertebral block (PVB), and intercostal nerve blocks (ICNB). Other techniques will be considered for inclusion on a case-by-case basis (with consensus of the screening authors). Each technique will be represented as an individual node within the treatment network.

#### Comparator

The comparators are either placebo, a passive comparison (e.g. standard care management with analgesia), or an active comparison (e.g. one block against another – head-to-head trials). Catheter-based interventions and neuraxial techniques will not be included as comparators.

#### Outcomes

We will include studies that have collected data any of the outcomes specified for this review (*see below*).

#### Study types

This review will include randomized clinical trials reported as published studies or abstracts from scientific congresses. There will be no restriction on publication status, language or year of publication.

#### Information sources

We will perform a search the electronic databases PubMed, MEDLINE, EMBASE, CinAHL and CENTRAL (the Cochrane Central Register of Controlled Trials) to identify published trials and conference abstracts. We will search clinical trial registries through the World Health Organisation international clinical trials registry platform and ClinicalTrials.gov. We will also search for unpublished trials through the PubMed indexed pre-print servers and by contacting primary authors of these studies. Reference lists of included studies plus other systematic reviews and meta-analyses will be manually reviewed.

#### Search strategy

We will develop our search strategy in alignment with the Peer Review of Electronic Search Strategies (PRESS) guideline statement^34^. Our search strategy will use a combination of search terms^35,36^ to identify patients with traumatic rib fractures and regional anaesthesia techniques (including by not limited to serratus anterior plane block (SAPB), erector spinae block (ESB), paravertebral block (PVB), and intercostal nerve blocks (ICNB)). The full electronic search strategy is shown in the Supplement.

#### Data management and selection process

Study records identified by our search strategy will be downloaded into COVIDENCE^37^. Duplicate records will be removed. Two authors will independently screen the titles and abstracts of these identified studies for their potential eligibility. Any differences or disagreements will be resolved by discussion or through a third reviewer. Reasons for exclusion will be recorded and presented in a Preferred Reporting Items for Systematic Reviews and Meta-analyses (PRISMA) flow diagram.

#### Data extraction process

A standardised digital data extraction sheet will be used through COVIDENCE. Two authors (CP and SF) will independently extract study information (see data items below) including information relevant to risk of bias grading. Any discrepancies will be resolved by discussion or by a third reviewer. If required, we will contact authors for further essential information, and cross check data against existing secondary sources.

#### Data items

We will extract data regarding study characteristics (including first author, year of publication, unit of randomisation, study type, number of participants, number of sites), details of the clinical setting (e.g., clinical speciality performing block, level of training of clinician, description of training program, ultrasound utilisation), participants (including age and sex distribution, mechanism of injury, concomitant injuries, number of ribs fractured, Anatomical Injury Severity [AIS] of thorax), interventions (regional technique(s) used, local anaesthetic used, dose administered, volume of diluent, and timing of technique, as well as details of the comparison group (alternate regional technique or usual care, co-intervention, intensive care admission, rib fixation surgery). Data related to primary and secondary outcomes (including timing of measurements) will also be collected as outlined below.

#### Outcomes

##### Primary outcome

The primary outcome will be pain, measured (as a representative score out of 10) using any of the following methods: visual analogue scale, numerical rating scale, PAINAD (for patients with dementia)^38^ at a time interval 4 to 8 hours post-block.

##### Secondary outcomes

The secondary outcomes will be;

1. Pain scores (as measured above) measured at 8-12 hours and at 24 hours, post-block.
2. Pain score reduction from baseline (reported as mean pain reduction).
3. Opioid requirements: Morphine milligram equivalents (MME)
  - Studies reporting doses of various individual opioid medications will have their doses converted to MME^39^.
4. Respiratory function
  - Measured by spirometry
  - Comparable score of peak flow, FEV1 (forced expiratory volume in one second), and FVC (forced vital capacity) will be used where able. Only likewise measures will be directly compared.
5. Diaphragmatic excursion (measured on ultrasound).
6. Respiratory complications: Pneumonia
7. Respiratory complications: Respiratory failure requiring non-invasive or invasive ventilation.
8. Procedural complication rate: Pneumothorax (PTX).
9. Procedural complication rate: Local anaesthetic systemic toxicity (LAST).
10. Hospital length of stay (days).
11. Mortality.

#### Risk of bias assessment

Two authors (CP and SF) will independently assess the methodological quality of the included studies for risk of bias. To assess risk of bias, we will use all publicly available reports of trials, including published trial protocols and statistical analysis plans. Any disagreements will be resolved by a senior review author (AD or KC). We will use the McMaster University CLARITY Group ‘Tool to Assess Risk of Bias in Randomized Controlled Trials’^40^. We will present a ‘risk of bias summary’ figure, depicting each risk of bias item for each included study (red high risk, green low risk, yellow unclear).

#### Data analyses

The main analyses will be performed on all included trials regardless of risk of bias. Studies will be pooled with traditional random-effects meta-analysis for each pairwise comparison. Effect sizes will be expressed as either relative risk (for dichotomous data) or weighted (or standardized) mean differences (for continuous data), and their 95% confidence intervals will be calculated for analysis. For pain score and pain score reductions when reported as median and interquartile range (IQR), mean and standard deviation will be calculated according to the formula reported by Wan *et al*^*41*^. Heterogeneity will be assessed statistically using the I^2^ test. A rigorous evaluation of clinical and methodological heterogeneity across the studies will provide information to make sure that the network maintains transitivity. Thus, a table of trial characteristics that may act as effect modifiers will be compiled from the data collected including patient demographics, concomitant injuries, and components of control arm (oral vs parenteral opioid, timing of block administration, allied health consultation).

If there is sufficient data, network meta-analysis will be conducted using a Bayesian approach to simultaneously compare multiple treatments via a common comparator (standard care)^42^. A network of treatment comparisons combining both direct and indirect evidence in a single model will be performed. To verify whether the information of both sources between direct and indirect evidence are similar enough to be combined (consistency assumptions), we will examine the regression coefficient of the inconsistency model for each study design and then test the linearity of the regression coefficients for all models. To explore statistical heterogeneity, sensitivity analyses will be conducted to assess the influence of the effect modifiers in the network. The ranking probabilities will be estimated based on the frequentist analogue of the surface under the cumulative ranking curve. Publication bias will be checked using funnel plots and Egger and Begg tests as long as more than 10 studies are included in the meta-analysis. All analyses will be implemented using the ‘‘meta’’, ‘‘netmeta’’, and “gemtc” packages of the R Environment for Statistical Computing v4.4.2 (Vienna, Austria)^43^.

A narrative summary will be provided if statistical analysis is not possible.

#### Subgroup analyses

Where sufficient data is available, we will conduct subgroup analyses for each outcome based on local anaesthetic agent. The credibility of any subgroup analysis will be assessed using the Instrument to assess the Credibility of Effect Modification Analyses (ICEMAN) in randomized clinical trials and meta-analyses^44^.

## Data Availability

All data produced as a result of this work will be contained in the subsequent manuscript.

## Confidence in cumulative evidence

The Grading of Recommendations Assessment, Development, and Evaluation (GRADE) approach will be used to evaluate the overall quality of evidence for each outcome measures^45,46^. Findings will be presented in a “Summary of findings and certainty of evidence” table. The certainty of evidence will be assessed based on five domains including the risk of bias, imprecision, inconsistency, indirectness and publication bias. For each outcome, the quality and certainty of evidence will be rated as “high”, “moderate”, “low” or “very low”.

## Supplement: Intended search strategies

### PUBMED

(((“clinical”[Title/Abstract] AND “trial”[Title/Abstract]) OR “clinical trials as topic”[MeSH Terms] OR “clinical trial”[Publication Type] OR “random*”[Title/Abstract] OR “random allocation”[MeSH Terms] OR “therapeutic use”[MeSH Subheading]) AND ((chest trauma) OR (rib fractures))) AND (((((((nerve block) OR (peripheral nerve block)) OR (pain management)) OR (serratus anterior)) OR (erector spinae)) OR (paravertebral)) OR (intercostal nerve)).

### MEDLINE (Ovid) (1946-present)

**Database: Ovid MEDLINE(R) ALL <1946 to present>**

**Search Strategy:**

1. Rib Fractures/ (3915)
2. rib fracture*.mp. (6132)
3. Thoracic Injuries/ (14017)
4. chest trauma.mp. (4533)
5. 1 or 2 or 3 or 4 (20742)
6. Nerve Block/ (22952)
7. nerve block*.mp. (34733)
8. serratus anterior.mp. or SAPB.tw. (2054)
9. erector spinae.mp. or ESB.tw. (5379)
10. paravertebral.mp. or PVB.tw. (7529)
11. intercostal nerve.mp. or ICNB.tw. (1550)
12. Pain Management/ (44271)
13. or 7 or 8 or 9 or 10 or 11 or 12 (88629)
14. 5 and 13 (494)

### EMBASE (Ovid) (1947-present)

**Database: Embase Classic+Embase <1947 to 2024 December 13>**

**Search Strategy:**

1. rib fracture/ (12027)
2. rib fracture*.mp. (13025)
3. 1 or 2 (13025)
4. nerve block/ (44839)
5. nerve block*.mp. (52243)
6. analgesia/ (173297)
7. serratus anterior.mp. or SAPB.tw. (3000)
8. erector spinae.mp. or ESB.tw. (7452)
9. paravertebral.mp. or PVB.tw. (12090)
10. intercostal nerve.mp. or ICNB.tw. (3940)
11. 4 or 5 or 6 or 7 or 8 or 9 or 10 (233358)
12. 3 and 11 (1236)
13. limit 12 to (clinical trial or randomized controlled trial or controlled clinical trial) (93)

### CINAHL Complete (via EBSCOhost)

CINAHL Complete (via EBSCOhost).
| # | Query | Results |
| --- | --- | --- |
| S12 | S4 AND S11 | 212 |
| S11 | S5 OR S6 OR S7 OR S8 OR S9 OR S10 | 52,178 |
| S10 | (MM "Pain Management") AND "pain management" | 39,398 |
| S9 | "intercostal nerve" | 361 |
| S8 | "paravertebral" | 1,547 |
| S7 | "erector spinae" | 2,209 |
| S6 | "serratus anterior plane" | 133 |
| S5 | (MH "Nerve Block+") AND "nerve block" | 12,076 |
| S4 | S1 OR S2 OR S3 | 5,495 |
| S3 | (MH "Thoracic Injuries+") AND "thoracic trauma" | 4,904 |
| S2 | "chest trauma" | 2,660 |
| S1 | (MM "Rib Fractures") AND "rib fracture" | 1,428 |

### CENTRAL

**Database: EBM Reviews - Cochrane Central Register of Controlled Trials <November 2024> Search Strategy:**

1. rib fractures.mp. [mp=title, original title, abstract, floating sub-heading word, mesh headings, heading words, keyword] (382)
2. thoracic injury.mp. [mp=title, original title, abstract, floating sub-heading word, mesh headings, heading words, keyword] (29)
3. chest trauma.mp. [mp=title, original title, abstract, floating sub-heading word, mesh headings, heading words, keyword] (186)
4. 1 or 2 or 3 (546)
5. nerve block.mp. [mp=title, original title, abstract, floating sub-heading word, mesh headings, heading words, keyword] (14805)
6. serratus anterior plane.mp. [mp=title, original title, abstract, floating sub-heading word, mesh headings, heading words, keyword] (517)
7. erector spinae.mp. [mp=title, original title, abstract, floating sub-heading word, mesh headings, heading words, keyword] (2827)
8. paravertebral.mp. [mp=title, original title, abstract, floating sub-heading word, mesh headings, heading words, keyword] (2501)
9. intercostal nerve.mp. [mp=title, original title, abstract, floating sub-heading word, mesh headings, heading words, keyword] (646)
10. pain management.mp. [mp=title, original title, abstract, floating sub-heading word, mesh headings, heading words, keyword] (16988)
11. 5 or 6 or 7 or 8 or 9 or 10 (33448)
12. 4 and 11 (184)

